# Real-time forecasting of measles transmission in Mexican states hosting FIFA World Cup venues, 2026

**DOI:** 10.64898/2026.02.17.26346510

**Authors:** Raj Kumar Subedi, Aida Jiménez Corona, Imelda Trejo Lorenzo, Hiroshi Nishiura, Isaac Chun-Hai Fung, Guillermo Carbajal-Sandoval, Miguel Ángel Nakamura-López, Maria Eugenia Jiménez Corona, Gerardo Chowell

## Abstract

Mexico has reported 18,092 confirmed cases of measles from January 2025-June 2026, shortly before the 2026 FIFA World Cup matches in Guadalajara, Mexico City and Monterrey. Using national surveillance data and the Generalized Richards Model, we generated 6-week forecasts for the two highest-burden venue states. Jalisco was projected to report 118 cases (95% PI: 5-427) and Ciudad de México 22 (95% PI: 0-89). Pre-departure check of MMR vaccination status and enhanced event-associated surveillance are warranted.

## Background and aim

Measles remains an important vaccine-preventable disease, and mass gatherings such as the FIFA World Cup can amplify transmission through crowding and mixing of populations with heterogeneous immunity profiles [1, 2]. Mexico co-hosts the 2026 FIFA World Cup, with matches in Guadalajara (Jalisco), Mexico City (Ciudad de México) and Monterrey (Nuevo León) from June 11 through July 19, 2026 [3], involving international travellers relevant to cross-border preparedness and travel-health messaging [4].

Since January 2025, Mexico has reported 18,092 confirmed measles cases across all 32 states in two temporally distinct waves, disproportionately affecting unvaccinated individuals [5]. We applied the Generalized Richards Model [6] to generate 6-week ahead forecasts for the two highest-burden World Cup venue states.

### The outbreak and transmission dynamics

We used confirmed case data from Mexico’s Dirección General de Epidemiología (DGE) through the Sistema Especial de Vigilancia Epidemiológica de Enfermedades Febriles Exantemáticas (EFE), with a data extract dated June 1, 2026. Cases were confirmed by laboratory testing (IgM serology or PCR; 96.5%) or clinico-epidemiologic criteria. Weekly incidence series were constructed by aggregating confirmed cases by epidemiological week of rash onset. To account for right-censoring due to reporting delays (median onset-to-classification: 5 days, IQR: 3-7), the final two weeks of each series were excluded from model calibration.

### Epidemic curve and reproduction number

We constructed the national weekly epidemic curve from onset dates of rash spanning epidemiological weeks 1-52 of 2025 and weeks 1-22 of 2026 (January 2025 - May 2026). The time-varying reproduction number *R*_*t*_ was estimated from symptom onset counts using a 7-day sliding window using the EpiEstim framework [7] with a parametric gamma-distributed serial interval (mean: 11.7 days, SD: 2.0 days) [8].

### State-level growth model forecasts

State-level 6-week ahead forecasts for Jalisco and Ciudad de México were generated using the Generalized Richards Model (GRM) [6]:

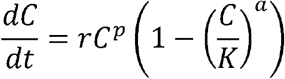

where *C*(*t*) is the cumulative case count, *r* is the intrinsic growth rate, *p* ∈ (0,1] controls early epidemic growth dynamics, *K* is the carrying capacity, and *a* > 0 controls epidemic curve asymmetry. Incidence at time *t* is *I*(*t*) = *dC*/*dt*.

Models were fitted by maximum likelihood estimation under a negative binomial error structure:

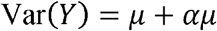

where *μ* is the model-predicted mean incidence and *α* > 0 is the overdispersion parameter. Parameter uncertainty was characterised using 300 parametric bootstrap replicates. Forecast prediction intervals (PI) represent the 2.5th and 97.5th percentiles of the bootstrap distribution. The initial condition *I*_O_ was fixed to the first observed value in each series. Calibration lengths were 37 weeks for Jalisco (epidemiological weeks 8-43, 2025) and 26 weeks for Ciudad de México (epidemiological week 45, 2025 - week 18, 2026). The forecast horizon was 6 weeks from the last calibration week (∼May 25, 2026), covering the full World Cup period. Week indices in the GrowthPredict toolbox plots are sequential from the first week of calibration and do not correspond directly to calendar epidemiological week numbers.

All analyses were conducted in MATLAB using the GrowthPredict toolbox [6] and R version 4.4.2 [9].

### National outbreak dynamics

From weeks 1-69, 2025-2026, Mexico reported 17,869 confirmed cases of measles from the June 1, 2026 data extract; the official DGE count of 18,092 as of June 5, 2026 reflects an additional 223 cases reported in the intervening four days [5]. The outbreak exhibited a bimodal pattern (Figure 1A). The first wave, concentrated in Chihuahua, peaked at 322 cases/week (epidemiological week 13, 2025) and declined to near-zero by epidemiological week 42, 2025; the second wave, driven predominantly by Jalisco, commenced in epidemiological week 47, 2025, peaked at 1,167 cases/week (epidemiological week 7, 2026), and was declining at the data extract (99 cases/week, epidemiological week 21). Jalisco contributed 39.1% of confirmed cases, followed by Chihuahua (25.9%) and Chiapas (6.3%). *R*_*t*_ peaked at 15.0 (95% CrI: 4.9-30.7) in February 2025 (Figure 1B), remaining above 1 nationally through February 2026 before declining below 1 at the time of the data extract. Among confirmed cases, 84.9% were unvaccinated, 10.3% had received one dose, and 4.9% two or more doses. The hospitalization rate was 14.7%, highest in children aged 1-4 years (29.7%).

**Figure 1.**
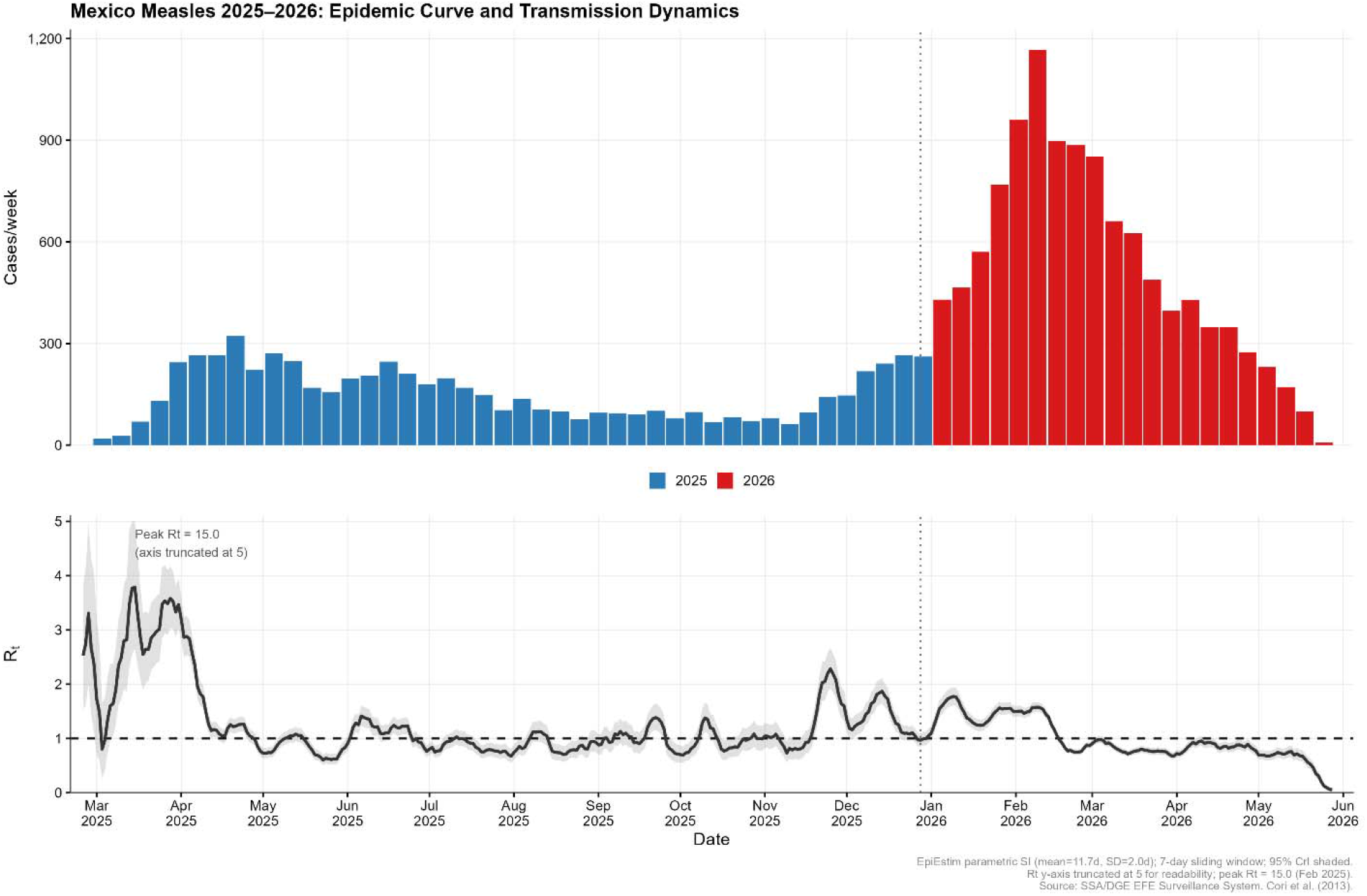
National measles epidemic curve and time-varying reproduction number (*R*_*t*_), Mexico, 2025–2026. Panel A: Weekly confirmed cases by epidemiological week of rash onset (blue: 2025; red: 2026). Dotted vertical line separates calendar years. Panel B: Time-varying *R*_*t*_(black line) with 95% credible interval (grey shading), estimated from symptom onset counts using a 7-day sliding window. Dashed horizontal line at *R*_*t*_ = 1. Y-axis truncated at 5 for readability; peak *R*_*t*_ = 15.0 (February 2025). Source: SSA/DGE EFE Surveillance System [5].

### State-level forecasts and World Cup host venues

Jalisco, with 6,980 total confirmed cases and 203 cases in the four weeks prior to the data extract, showed a declining trajectory at forecast origin (Figure 2, Panel A). The GRM projected a mean of 118 cases (95% PI: 5-427) in Jalisco during the World Cup period (Table 1). Ciudad de México reported 44 cases in the four weeks prior to the data extract and is projected to report a mean of 22 cases (95% PI: 0-89) (Figure 2, panel B). Monterrey (Nuevo León) reported 9 cases in the four weeks prior to the data extract, indicating low current transmission (Table 1; Figure 3).

**Table 1.** Recent transmission intensity at World Cup venue states, Mexico, 2026.

| Venue | State | Cases (last 4 weeks)<sup>a</sup> | 6-week forecast, mean (95% PI)<sup>b</sup> | Intensity of transmission |
| --- | --- | --- | --- | --- |
| Estadio Akron, Guadalajara | Jalisco | 203 | 118 (5-427) | High |
| Estadio Azteca / Estadio Ciudad de México, Mexico City | Ciudad de México | 44 | 22 (0-89) | Moderate |
| Estadio BBVA, Monterrey | Nuevo León | 9 | - | Low |

<sup>a</sup> Confirmed cases in the four weeks prior to the data extract (June 1, 2026).
<sup>b</sup> State-level cumulative 6-week forecast (~May 26-July 13, 2026) derived using the Generalized Richards Model with negative binomial error and 300 bootstrap replicates; PI: prediction interval.

**Figure 2.**
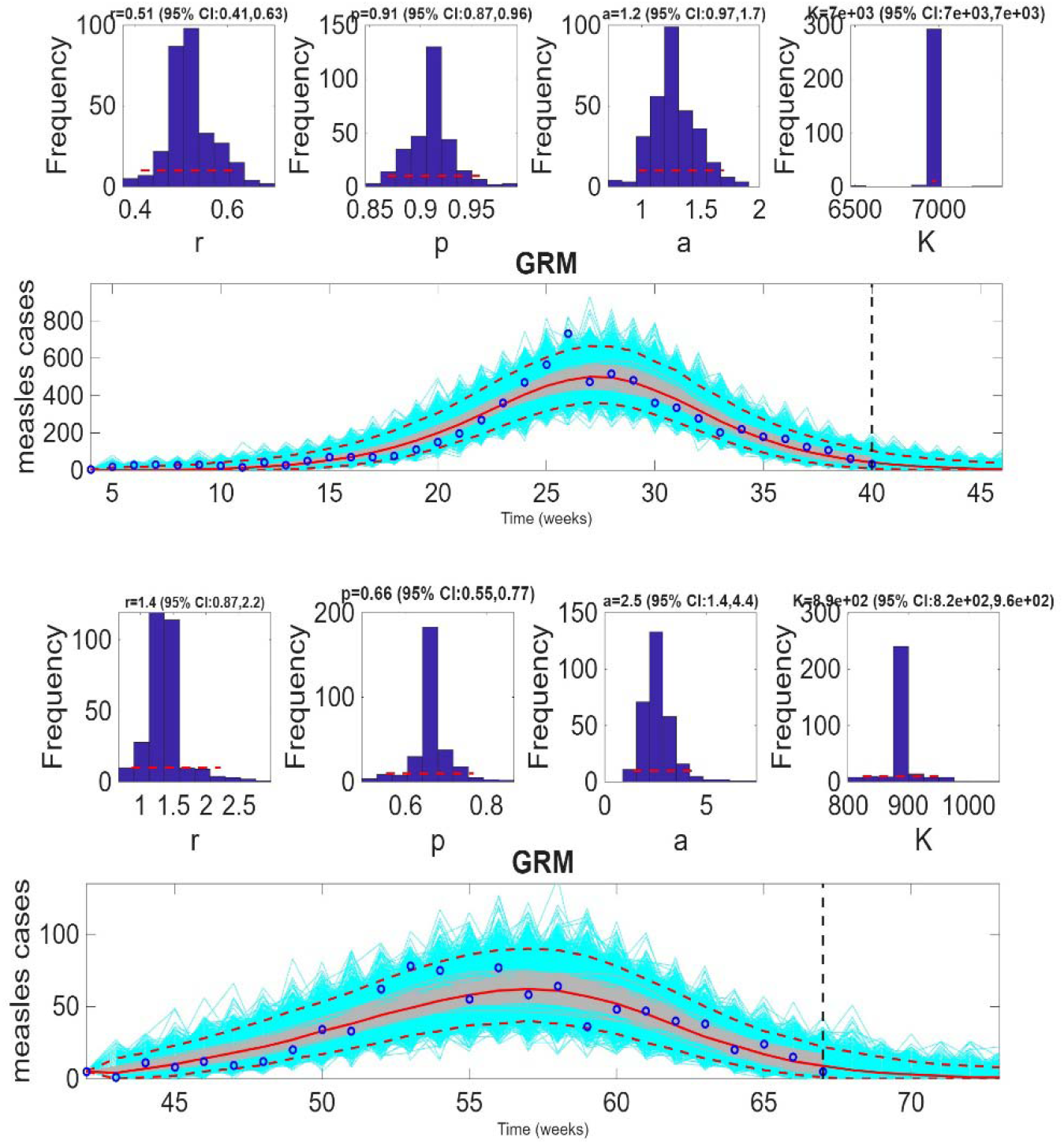
State-level Generalized Richards Model fit and 6-week ahead forecast, World Cup venue states, Mexico, 2026. Blue circles: observed weekly confirmed cases. Red line: GRM mean fit. Grey shading: 95% bootstrap prediction interval. Dashed vertical line: forecast origin (∼May 25, 2026). Panel A-Jalisco: calibration epidemiological weeks 8-43 (*n* = 37; *r* = 0.5, p= 0.9, *a* = 1.2, *K* = 6,980). Panel B-Ciudad de México: calibration epidemiological week 45, 2025 - week 18, 2026 (*n* = 26; *r* = 1.4, *p* = 0.7, *a* = 2.5, *K* = 891). Forecasts represent state-level totals; prediction intervals reflect parameter uncertainty under 300 bootstrap replicates. Source: SSA/DGE EFE Surveillance System [5].

**Figure 3.**
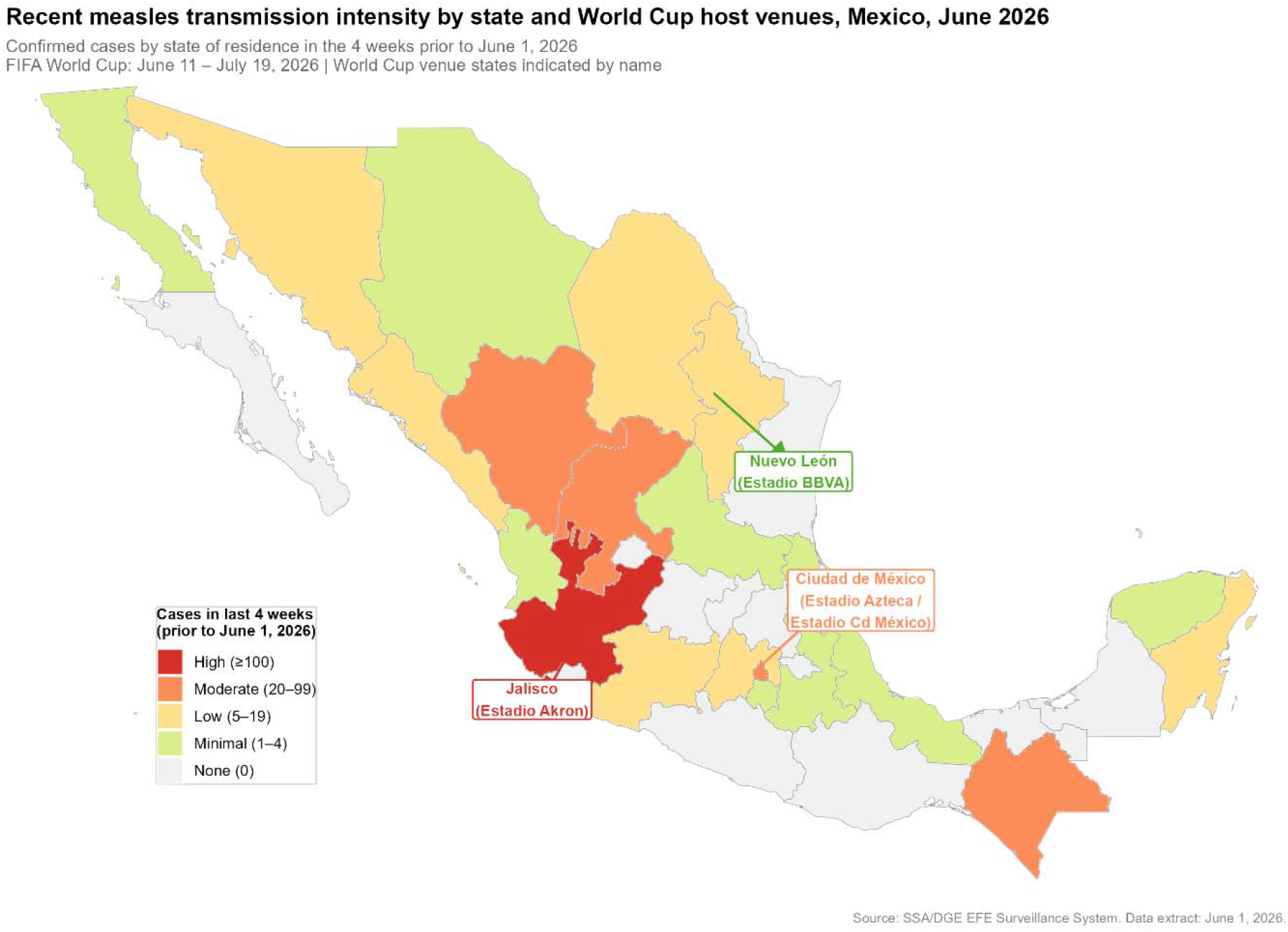
Recent transmission intensity by state and World Cup host venues, Mexico, June 2026. States shaded by confirmed cases in the four weeks prior to June 1, 2026 (intensity categories: High ≥100, Moderate 20-99, Low 5-19, Minimal 1-4, None 0). World Cup venue states are indicated by name. Source: SSA/DGE EFE Surveillance System [5].

## Discussion

We present short-term measles transmission forecasts for the 2026 FIFA World Cup venue states in Mexico [3], in the context of an ongoing national outbreak. Our findings suggest that transmission is most active in Jalisco, where 203 cases were reported in the four weeks prior to the data extract and the 6-week state-level forecast projects a mean of 118 cases (95% PI: 5-427) during the tournament. The wide prediction interval reflects genuine uncertainty about the pace of ongoing decline, with the lower bound suggesting transmission could approach near-zero and the upper bound indicating that some level of continued activity of transmission remains plausible. Ciudad de México (Mexico City) shows moderate recent transmission with a projected mean of 22 cases (95% PI: 0-89), and Monterrey shows minimal activity. Age-stratified forecasting was beyond the scope of this analysis; future work incorporating age-specific vaccination coverage data could provide more refined transmission estimates across population groups.

The findings support targeted preparedness rather than alarm. However, mass gathering-related mobility, crowding and mixing of susceptible travelers could temporarily increase transmission even though incidence has been declining. Before travel departure, spectators, teams and staff should verify that they have completed age-appropriate measles-mumps-rubella (MMR) vaccination, particularly those travelling to or from Jalisco and Ciudad de México. ECDC notes that all EU/EEA countries include two MMR doses in their national immunisation schedules and that two doses are needed for full protection against measles [4].

During and after the tournament, host and origin-country public health authorities should maintain awareness for fever-rash illness among travellers, ensure rapid isolation and notification of suspected measles cases, and support contact tracing and event-associated surveillance. Given that many cases in North America occurred among unvaccinated people, awareness communication should emphasise timely MMR vaccination. Because tournament-related exposures may result in rash onset after the 6-week forecast horizon, post-travel surveillance should continue after the event.

We acknowledge some limitations. Forecasts are based on data with a 7-day reporting lag; incidence in the final calibration weeks may be slightly underestimated due to ongoing case classification. The GRM does not incorporate population mixing dynamics specific to the World Cup or potential vaccination campaigns during the forecast period, either of which could shift the trajectory. The wide prediction intervals, particularly for Jalisco, underscore the inherent uncertainty in short-term epidemic forecasting and should be interpreted accordingly.

## Conclusion

Measles transmission remains active in Mexico, with Jalisco representing the highest-burden World Cup venue state. Short-term forecasts suggest continued but declining transmission during the tournament period. Nevertheless, mass-gathering-related mobility and mixing of susceptible people could temporarily increase transmission, even during a declining phase. Pre-travel confirmation of age-appropriate MMR vaccination, prompt detection and isolation of suspected cases, and routine event-associated surveillance are recommended as part of World Cup health preparedness planning.

## Data Availability

All data produced are available online at https://www.gob.mx/salud/documentos/informe-diario-del-brote-de-sarampion-en-mexico-2026

## Acknowledgement

This work was partly supported by UNAM through the PREI-DGAPA. The authors acknowledge the collaboration with the host institution, Centro de Ciencias Matemáticas, Universidad Nacional Autónoma de México, during G.C’s research stay. The authors thank the General Directorate of Epidemiology for facilitating the data of the National Epidemiological Surveillance System.

